# Association Between Blood Cadmium Levels and Stroke Risk: A Cross-sectional Analysis of NHANES 1999-2018

**DOI:** 10.1101/2025.06.23.25330115

**Authors:** Baohao Zhu, Debiao Yu, Peng Chen, Wanzhu Wang, Fangyuan Xu, Xi Li, Long Huang, Bin Shao

## Abstract

**Background:** Stroke is a leading cause of mortality and disability worldwide, with traditional risk factors explaining only 60-70% of stroke pathogenesis. The association between cadmium exposure, an emerging environmental toxin, and stroke risk remains unclear, particularly regarding the potential mediating role of inflammatory responses. This study aimed to systematically evaluate the association between blood cadmium concentrations and stroke prevalence using data from the National Health and Nutrition Examination Survey (NHANES) 1999-2018.

**Methods:** This cross-sectional study analyzed 10,434 participants aged ≥20 years from NHANES 1999-2018. Blood cadmium levels were measured using inductively coupled plasma mass spectrometry. Stroke prevalence was assessed through structured interviews. Multivariable logistic regression, restricted cubic splines, and mediation analysis were employed to evaluate associations and potential biological pathways.

**Results:** Among 10,434 participants, 404 (3.9%) had a history of stroke. Blood cadmium concentrations were significantly higher in stroke patients compared to non-stroke individuals (0.66±0.68 μg/L vs 0.52±0.61 μg/L, P<0.001). After adjusting for demographics, lifestyle factors, and comorbidities, each 1 μg/L increase in blood cadmium was associated with a 48.8% increased stroke risk (OR=1.488, 95% CI: 1.292-1.713, P<0.001). The highest cadmium quartile showed a 118.7% increased stroke risk compared to the lowest quartile (OR=2.187, 95% CI: 1.413-3.384, P<0.001). Mediation analysis revealed that white blood cell count mediated 11.0% of the association (95% CI: 2.1%, 19.9%), suggesting inflammatory pathways as potential mechanisms.

**Conclusions:** Blood cadmium exposure is significantly associated with increased stroke risk, with evidence of a clear dose-response relationship. Inflammatory mechanisms may partially mediate this association. These findings highlight the importance of environmental cadmium exposure as a modifiable risk factor for stroke prevention.

## Introduction

Stroke represents one of the leading causes of mortality and disability worldwide[1]. According to the latest Global Burden of Disease Study, approximately 13 million new stroke cases occur annually, imposing a substantial burden on global public health systems[2]. Despite decades of research, traditional risk factors including hypertension, diabetes mellitus, dyslipidemia, and smoking account for only 60-70% of stroke pathogenesis[3], leaving a substantial proportion of stroke etiology unexplained. This knowledge gap has prompted increasing attention toward environmental factors, particularly heavy metal exposures, as potential contributors to the remaining stroke risk[4].

Among environmental toxicants, cadmium (Cd) has emerged as a particularly concerning cardiovascular risk factor due to its unique bioaccumulation characteristics and widespread environmental distribution[5, 6]. Cadmium exposure occurs primarily through smoking, dietary intake, industrial emissions, and occupational exposure[7–9]. Notably, a recent scientific statement by the American Heart Association emphasized cadmium as an established risk factor for cardiovascular diseases, yet clinical recognition and targeted interventions remain underdeveloped[10]. Meta-analyses demonstrate that cadmium exposure increases stroke risk by up to 85, highlighting its potential clinical significance%[5, 11]. However, existing studies show inconsistent results regarding cadmium-stroke associations, possibly due to differences in study populations, exposure assessment methods, and confounding factor control[12, 13].

The biological mechanisms underlying cadmium-induced stroke risk appear to converge on pathways central to stroke pathogenesis itself[14]. Cadmium exposure activates inflammatory signaling pathways, upregulating pro-inflammatory cytokines such as tumor necrosis factor-α and interleukin-6[15]. Simultaneously, cadmium induces reactive oxygen species production in vascular endothelial cells, enhancing oxidative stress responses and disrupting vascular endothelial barrier function[16]. These pathological processes—inflammation and oxidative stress—are also fundamental mechanisms in stroke development, suggesting that cadmium may exacerbate stroke risk through amplifying existing pathophysiological pathways[17].

Despite these established mechanistic links, several critical gaps remain in our understanding of cadmium-stroke relationships[18]. First, most existing studies have relied on single inflammatory biomarkers or focused on general cardiovascular outcomes rather than stroke specifically[19].

Second, the mediating role of inflammatory pathways in cadmium-stroke associations has not been systematically evaluated using causal mediation analysis[20]. Third, studies have largely overlooked potential population heterogeneity in cadmium susceptibility, particularly regarding metabolic status and racial differences[21]. Fourth, the dose-response relationship between cadmium exposure and stroke risk remains poorly characterized, with limited exploration of potential threshold effects or non-linear patterns[22].

Based on these considerations, we hypothesize that cadmium exposure significantly increases stroke risk through inflammatory-mediated pathways. The present study utilized data from the National Health and Nutrition Examination Survey (NHANES) 1999-2018 to systematically evaluate the association between blood cadmium concentrations and stroke prevalence, explore dose-response relationships, and assess the mediating role of inflammatory biomarkers. By integrating nearly 20 years of nationally representative data with advanced statistical methodologies including restricted cubic splines and causal mediation analysis, this study aims to provide definitive evidence for cadmium’s role in stroke etiology and inform targeted prevention strategies.

## 2 Materials and Methods

### 2.1 Study Design and Population

This study utilized data from the National Health and Nutrition Examination Survey (NHANES) 1999-2018 to investigate the association between blood cadmium concentrations and stroke incidence. NHANES is a comprehensive cross-sectional survey conducted biennially to assess the health and nutritional status of the non-institutionalized civilian US population. The survey strictly adheres to ethical standards established by the National Center for Health Statistics (NCHS) Research Ethics Review Board, with all participants required to provide informed consent. We excluded individuals with unclear stroke diagnoses, incomplete cadmium levels or covariate data, and those lacking important laboratory examination data. The final study sample included 10,434 subjects with national representativeness.(Fig. 1)

**Fig. 1.**
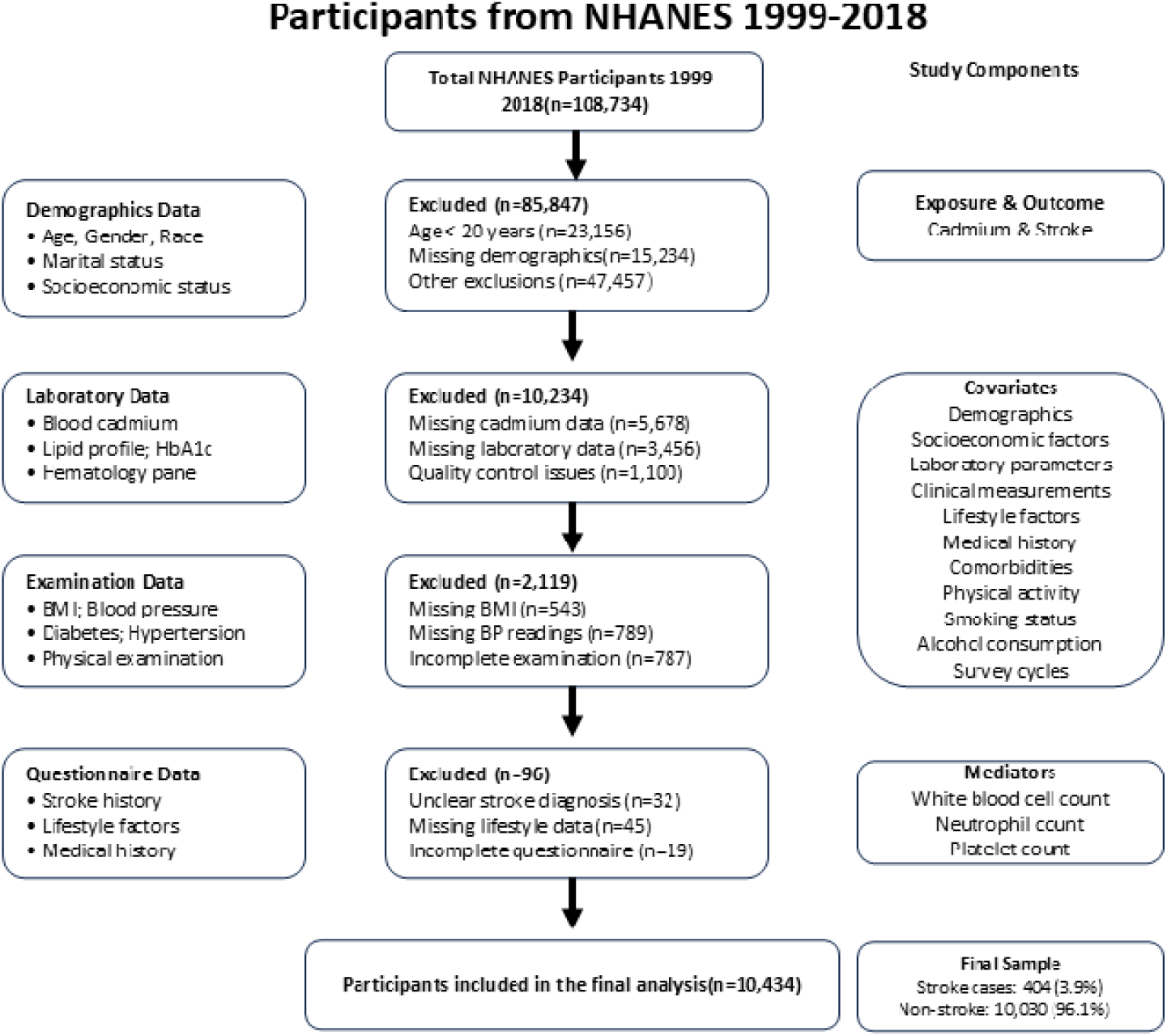
Flowchart of participants included in this study. NHANES, National Health and Nutrition Examination Survey; BMI, body mass index; BP, blood pressure; HbA1c, hemoglobin A1c.

### 2.2 Stroke Assessment

Stroke occurrence was collected through the structured interview process in the NHANES protocol. Researchers asked participants: "Have you ever been diagnosed with stroke by a healthcare professional?" In this study, affirmative responses were considered indicators of previous stroke events. Participants who answered "don’t know," "refused to answer," or did not respond to the question were excluded from analysis to ensure clarity and accuracy of stroke diagnosis data.

### 2.3 Cadmium Analysis

Blood specimens were preserved and processed according to NHANES standardized protocols, then sent to the Centers for Disease Control and Prevention (CDC/NCHS) Laboratory Sciences Division for analytical processing. Blood cadmium concentrations were quantitatively detected using inductively coupled plasma mass spectrometry. Considering that over 80% of measurements might be below the detection limit, NHANES employed the detection limit divided by √2 method to estimate concentrations below the detection limit, as described in the CDC’s “National Report on Human Exposure to Environmental Chemicals[23]." We categorized blood cadmium concentrations (LBXBCD) using quartiles (Q1-Q4) to study their association with stroke prevalence, with the lowest quartile (Q1) serving as the reference category.

### 2.4 Variable Definition and Classification

We considered a series of clinically relevant variables in our analysis, including age at interview (RIDAGEYR), gender (RIAGENDR), race/ethnicity (RIDRETH1), marital status (DMDMARTL), BMI classification (BMI_category), alcohol consumption (Alcohol_Consumption), smoking status (Smoking_Status), physical activity level (activity_level), diabetes (Diabetes), and hypertension history (Hypertension).

Age was recorded in years, and gender was classified as male and female. Race/ethnicity was categorized according to NHANES classification standards as Non-Hispanic White, Non-Hispanic Black, Mexican American, Other Hispanic, and Other Race-including multi-racial mixed populations. Marital status was classified into categories including married, widowed, separated, never married, and living together. BMI was classified into four categories: underweight, normal weight (reference category), overweight, and obese. Alcohol consumption was classified as drinkers and non-drinkers. Smoking status was categorized as never smoker, occasional smoker, former smoker, recent quitter, and long-term quitter. Physical activity level was classified into low and moderate levels.

Diabetes was determined through survey responses, current diabetes medication use, or meeting any of the following criteria: fasting glucose ≥126 mg/dL, 2-hour postprandial glucose ≥200 mg/dL, or glycated hemoglobin ≥6.5%. Hypertension was determined based on self-reported diagnosis, current antihypertensive medication use, or blood pressure readings ≥140/90 mmHg. Triglyceride levels were classified into three groups according to clinical standards: normal group (<150 mg/dL), borderline group (150-199 mg/dL), and high level group (≥200 mg/dL) for stratified analysis of stroke risk heterogeneity assessment.

### 2.5 Laboratory Parameters

Additional laboratory parameters were included to assess potential mediation pathways and confounding factors. Lipid profiles included total cholesterol (TC), triglycerides (TG), high-density lipoprotein cholesterol (HDL), and low-density lipoprotein cholesterol (LDL). Hematological parameters included hemoglobin (HB), red blood cell count (RBC), white blood cell count (WBC), neutrophil count (NEU), monocyte count (MONO), lymphocyte count (LYM), and platelet count (PLT). These variables were included to examine their potential roles as mediating factors or confounding factors in the relationship between cadmium exposure and stroke risk.

### 2.6 Stratified Analysis

To explore heterogeneity in the association between cadmium exposure and stroke risk across different population characteristics, we conducted systematic stratified analyses. Specifically, we examined the association strength and patterns between cadmium quartiles and stroke risk in different subgroups. Stratified analyses included stratification by blood lipid levels (triglyceride normal group <150 mg/dL, borderline group 150-199 mg/dL, high level group ≥200 mg/dL) and race/ethnicity (Mexican American, Other Hispanic, Non-Hispanic White, Non-Hispanic Black, Other Race). This stratified analysis approach helps identify population differences in cadmium exposure effects on stroke risk, providing evidence for individualized risk assessment.

### 2.7 Statistical Models and Covariates

This analysis employed three models to assess the association between blood cadmium concentrations and stroke risk. The crude model provided initial estimates without adjusting for confounding factors. Model I adjusted for demographic and socioeconomic factors, including age, gender, race/ethnicity, and marital status. These variables were selected because they are established stroke risk factors that might confound the association between cadmium exposure and stroke risk. Model II further adjusted for lifestyle factors, comorbidities, and physical activity levels, including BMI classification, diabetes, hypertension, smoking status, alcohol consumption, and physical activity level, to better control for potential confounding and mediating factors.

### 2.8 Statistical Analysis

Preliminary comparative analysis of baseline data employed appropriate statistical testing methods for continuous and categorical variables. Categorical data were presented as counts and percentages (n, %), while continuous data were presented as mean ± standard deviation (SD) or median and interquartile range (IQR) according to data distribution. Considering NHANES complex survey design, all analyses incorporated appropriate sample weights (WTMEC2YR_adjusted), clustering (SDMVPSU), and stratification (SDMVSTRA) variables according to NCHS guidelines.

Logistic regression models were used to assess the association between cadmium exposure and stroke prevalence, with the lowest quartile (Q1) as the reference category. Results were presented as odds ratios (OR) and 95% confidence intervals (CI). The analysis process included crude models, Model I adjusting for demographic factors, and fully adjusted Model II incorporating lifestyle and comorbidities. Trend tests were also conducted by including cadmium quartiles as continuous variables in models to assess dose-response relationships.

To assess potential non-linear relationships, restricted cubic spline (RCS) analysis was employed to explore dose-response relationships between cadmium exposure and stroke risk. RCS analysis was set with 3 knots, with knot positions determined according to blood cadmium concentration distribution percentiles, set at the 10th percentile, 50th percentile (median), and 90th percentile. The rationale for choosing 3 knots was based on the following considerations: (1) balancing model complexity and statistical power while avoiding overfitting; (2) following recommendations by Harrell et al., suggesting that 3-4 knots are usually sufficient to capture non-linear relationships for studies with sample sizes >10,000; (3) considering the relatively small number of stroke events (404 cases), limiting the number of knots helps maintain model stability; (4) preliminary exploratory analysis showed that increasing the number of knots did not significantly improve model fit.

RCS analysis was conducted in different subgroups, including analyses stratified by triglyceride levels and race/ethnicity. For each stratified analysis, we calculated the significance of overall association (P-overall) and non-linear association (P-non-linear) to identify potential effect modification and non-linear patterns. RCS analysis was implemented using the rcs() function in the rms package of R software, with non-linear relationship statistical significance tested through the anova() function.

Subgroup analyses were conducted using stratified analysis methods, independently fitting logistic regression models and restricted cubic spline models in each subgroup. Forest plots were used to display association strengths between cadmium exposure and stroke risk in different subgroups, assessing effect heterogeneity. Stratified analysis results were evaluated for clinical significance of differences by comparing effect sizes and confidence interval overlaps across different subgroups. Subgroups included age, gender, race/ethnicity, BMI categories, diabetes status, hypertension status, smoking status, alcohol consumption, and physical activity levels. Interaction tests were performed by adding interaction terms between cadmium exposure and stratification variables in logistic regression models, with P < 0.05 as the statistical significance criterion.

Mediation analysis was conducted to examine potential biological pathways connecting cadmium exposure and stroke risk. Mediation analysis employed a causal mediation analysis framework based on potential outcome model theory[24]. Specific analysis steps followed the classic Baron and Kenny method, combined with modern causal inference methods:(1) Establish total effect model: Y = α + c·X + β·C + εLJ, where Y is stroke outcome, X is cadmium exposure, C is covariate vector; (2) Establish mediator model: M = α + a·X + γ·C + εLJ, where M is potential mediator; (3) Establish direct effect model: Y = α + c’·X + b·M + δ·C + εLJ; (4) Calculate indirect effect: IE = a × b, total effect: TE = c, direct effect: DE = c’, mediation proportion: PM = IE/TE × 100%.We separately examined the roles of lipid parameters (HDL cholesterol) and hematological indicators (neutrophil count, platelet count, white blood cell count) as potential mediating factors. Mediation analysis was conducted using the mediation package in R software, employing quasi-Bayesian Monte Carlo methods to calculate mediation effects and their 95% confidence intervals, with simulation times set to 1000. Statistical significance of indirect effects was assessed through bootstrap methods, with mediation effects considered significant when 95% confidence intervals did not include 0.

To ensure robustness of mediation analysis results, we conducted the following sensitivity analyses: (1) adopting different covariate adjustment strategies; (2) using continuous and categorical cadmium exposure variables for analysis; (3) re-analyzing after excluding extreme values. Mediation analysis considered NHANES complex sampling design, using appropriate weight adjustments when calculating standard errors.

All statistical analyses were performed using R software (version 4.3.0), with main packages including: survey package for handling complex sampling design, rms package for restricted cubic spline analysis, mediation package for mediation analysis, and forestplot package for drawing forest plots. Statistical significance was set at two-tailed P value <0.05. Considering NHANES complex survey design, variance estimation procedures suitable for clustered and stratified sampling were employed throughout the analysis process, using Taylor series linearization methods to calculate standard errors.

## 3 Results

### 3.1 Demographic and Clinical Characteristics of the Study Cohort

Table 1 describes the demographic and clinical characteristics of 10,434 participants in the NHANES study from 1999-2018. In this cohort, 404 individuals (3.9%) were identified as having stroke. Comparative analysis showed significant differences between stroke and non-stroke groups across multiple parameters, including age, race/ethnicity, marital status, BMI classification, diabetes prevalence, hypertension incidence, smoking status, alcohol consumption, physical activity level, and blood cadmium concentrations (P < 0.05). The mean age of stroke patients was significantly higher than non-stroke individuals (67.66±13.38 years vs 49.81±16.80 years, P<0.001). Blood cadmium concentrations were significantly higher in the stroke group compared to the non-stroke group (0.66±0.68 μg/L vs 0.52±0.61 μg/L, P<0.001).

**Table1.**
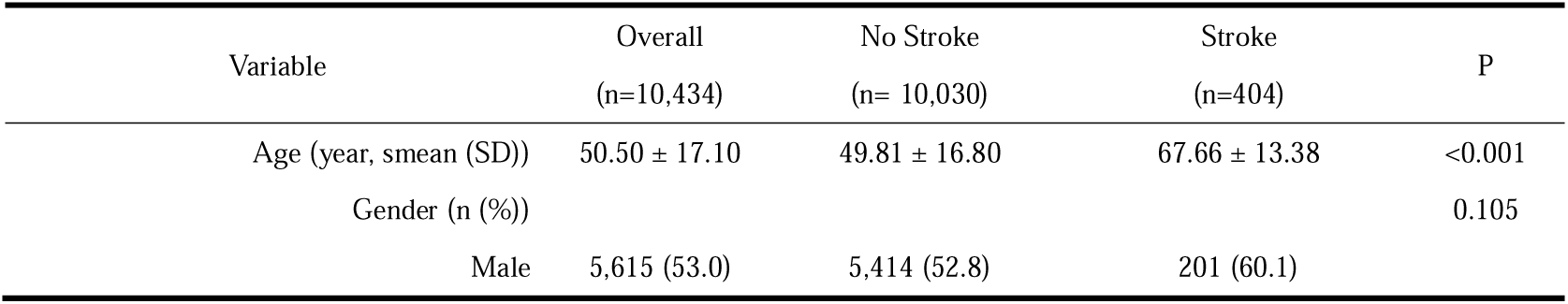
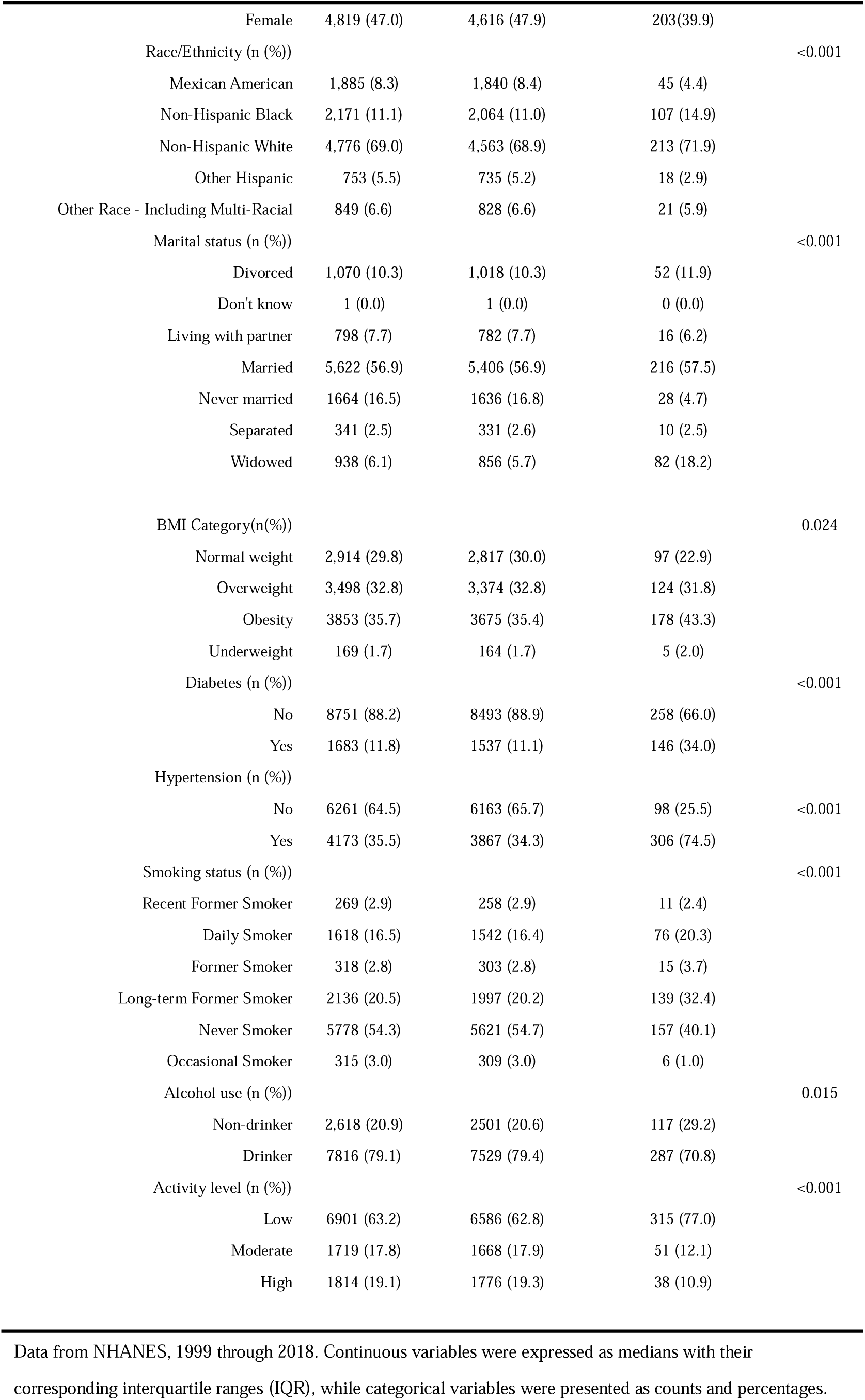
Baseline characteristics of study population by disease status.

### 3.2 Association Between Cadmium Exposure and Stroke Risk

The lowest quartile (Q1) was designated as the reference. Study results showed that in unadjusted analysis, elevated blood cadmium levels were associated with higher stroke prevalence, with Q4 compared to Q1 showing a crude OR of 3.412 (95% CI: 2.38-4.891, P<0.001) (Table 2). In adjusted Model I, after adjusting for age, gender, race/ethnicity, and marital status, cadmium maintained a positive correlation with stroke risk, with Q4 compared to Q1 showing an OR of 1.823 (95% CI: 1.217-2.731, P=0.005). Trend testing showed a significant association between blood cadmium concentration and stroke risk (trend OR=1.241, 95% CI: 1.077-1.431, P=0.004).

**Table2.** Association between blood cadmium quartiles and stroke risk: odds ratios from logistic regression models.

| Variable | Crude model |  | Model I |  | Model II |  |
| --- | --- | --- | --- | --- | --- | --- |
|  | OR (95%CI) | P | OR (95%CI) | P | OR (95%CI) | P |
| Q1 | 0.014 (0.01-0.018) | <0.001 | 0 (0-0.001) | <0.001 | 0 (0-0.002) | <0.001 |
| Q2 | 1.807<br>(1.245-2.623) | 0.002 | 1.032<br>(0.712-1.495) | 0.869 | 1.129<br>(0.766-1.664) | 0.542 |
| Q3 | 2.622 (1.843-3.73) | <0.001 | 1.069<br>(0.709-1.614) | 0.75 | 1.221<br>(0.771-1.933) | 0.397 |
| Q4 | 3.412 (2.38-4.891) | <0.001 | 1.823<br>(1.217-2.731) | 0.005 | 2.187<br>(1.413-3.384) | <0.001 |
| Trend | 1.472<br>(1.318-1.645) | <0.001 | 1.241<br>(1.077-1.431) | 0.004 | 1.313<br>(1.13-1.527) | <0.001 |

In Model II analysis, after further adjusting for BMI classification, diabetes, hypertension, alcohol consumption, and physical activity level, each 1 μg/L increase in blood cadmium concentration as a continuous variable increased stroke risk by 48.8% (OR=1.488, 95% CI: 1.292-1.713, P<0.001). Compared to the lowest quartile, the highest cadmium quartile was significantly associated with stroke risk (OR=2.187; 95% CI: 1.413-3.384, P<0.001).

To verify result robustness, we constructed multiple sensitivity analysis models with different adjustment strategies, including lipid parameters (Model II-B), physical activity (Model II-C), survey year (Model II-D), and hematological indicators (Model II-E). In all models not including smoking, cadmium exposure effects maintained statistical significance, with continuous variable ORs ranging from 1.480-1.543, Q4 compared to Q1 ORs ranging from 2.141-2.236, trend ORs ranging from 1.301-1.324, all P values <0.001. Linear trend analysis confirmed significant associations between log10-transformed cadmium concentrations and stroke prevalence in all multivariable regression models (trend OR=1.313, 95% CI: 1.130-1.527, P<0.001).

This study chose not to adjust for smoking status, primarily based on cadmium’s bioaccumulation characteristics and causal relationship considerations. Descriptive analysis showed significant differences in blood cadmium concentrations among different smoking status populations: daily smokers’ blood cadmium concentrations (1.309±0.987 μg/L) were nearly 4 times higher than never smokers (0.330±0.242 μg/L), and even in long-term quitters, blood cadmium concentrations (0.419±0.298 μg/L) remained significantly higher than never smokers. With smoking as a major pathway for cadmium exposure, adjusting for smoking might lead to over-adjustment bias, masking cadmium’s true health effects. Additionally, cadmium’s long biological half-life (10-30 years) makes its health impacts potentially persist independently of current smoking status.

### 3.3 Non-linear Analysis of Dose-Response Relationships

Restricted cubic spline (RCS) analysis further explored dose-response relationships between cadmium exposure and stroke risk. RCS analysis stratified by triglyceride levels showed different association patterns. In the normal triglyceride group (<150 mg/dL, N=7221), cadmium exposure showed a linear positive correlation with stroke risk (P = 0.008, P-non-linear = 0.39), indicating that in populations with normal blood lipids, increasing cadmium concentrations showed stable linear growth relationships with stroke risk.

In the borderline high triglyceride group (150-199 mg/dL, N=1434), no significant association was observed (P = 0.35, P-non-linear = 0.63), suggesting weakened associations between cadmium exposure and stroke risk in populations with mildly elevated triglycerides. In the high triglyceride group (≥200 mg/dL, N=1400), significant overall associations and non-linear relationships were observed (P = 0.002, P-non-linear = 0.039), showing complex dose-response patterns where risk peaked at moderate cadmium exposure concentrations, followed by slight decreases at higher concentrations. (Fig. 2)

**Fig. 2.**
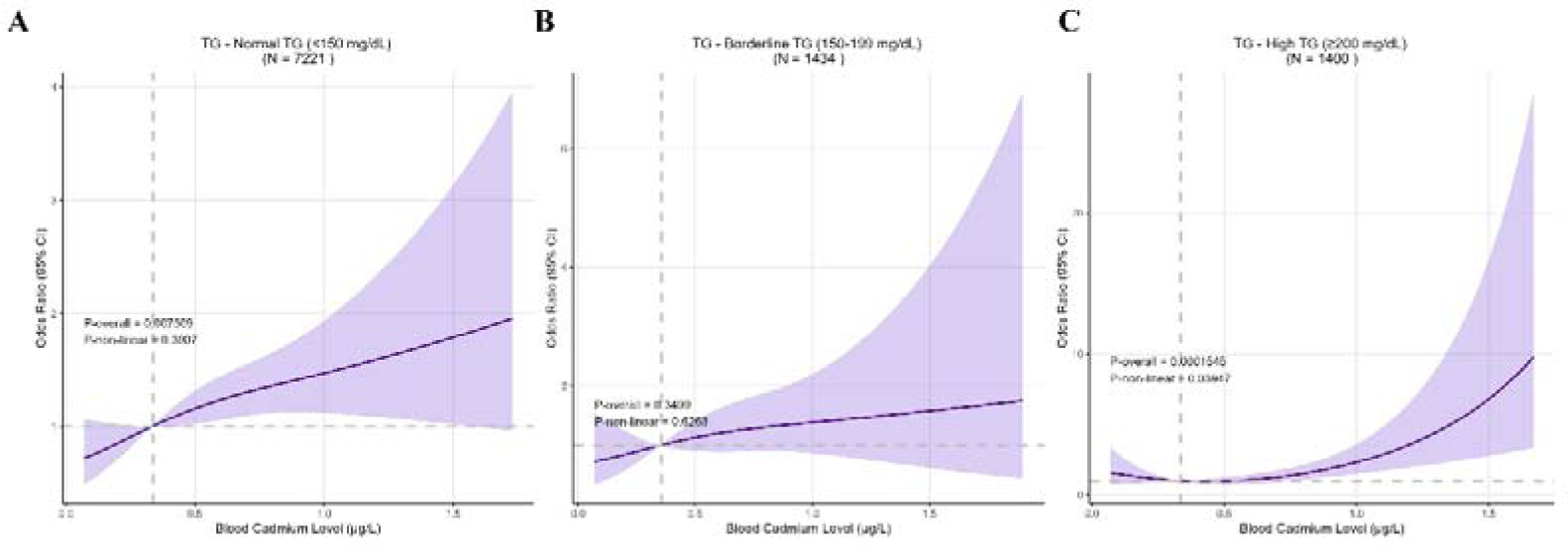
Restricted cubic splines showing dose-response relationships between blood cadmium levels and stroke risk by triglyceride status. (A) Normal TG (<150 mg/dL), (B) Borderline TG (150-199 mg/dL), (C) High TG (≥200 mg/dL). Solid lines indicate odds ratios with 95% confidence intervals (shaded areas). TG, triglycerides.

### 3.4 Dose-Response Analysis in Race/Ethnicity Subgroups

RCS analysis stratified by race/ethnicity showed racial differences in cadmium exposure effects on stroke risk. The strongest association was observed in Non-Hispanic Blacks (P = 0.003), followed by other racial groups (P < 0.001). Associations were weaker in Non-Hispanic Whites (P = 0.085). Associations in Mexican Americans and Other Hispanic groups did not reach statistical significance (P = 0.24). (Fig. 3)

**Fig. 3.**
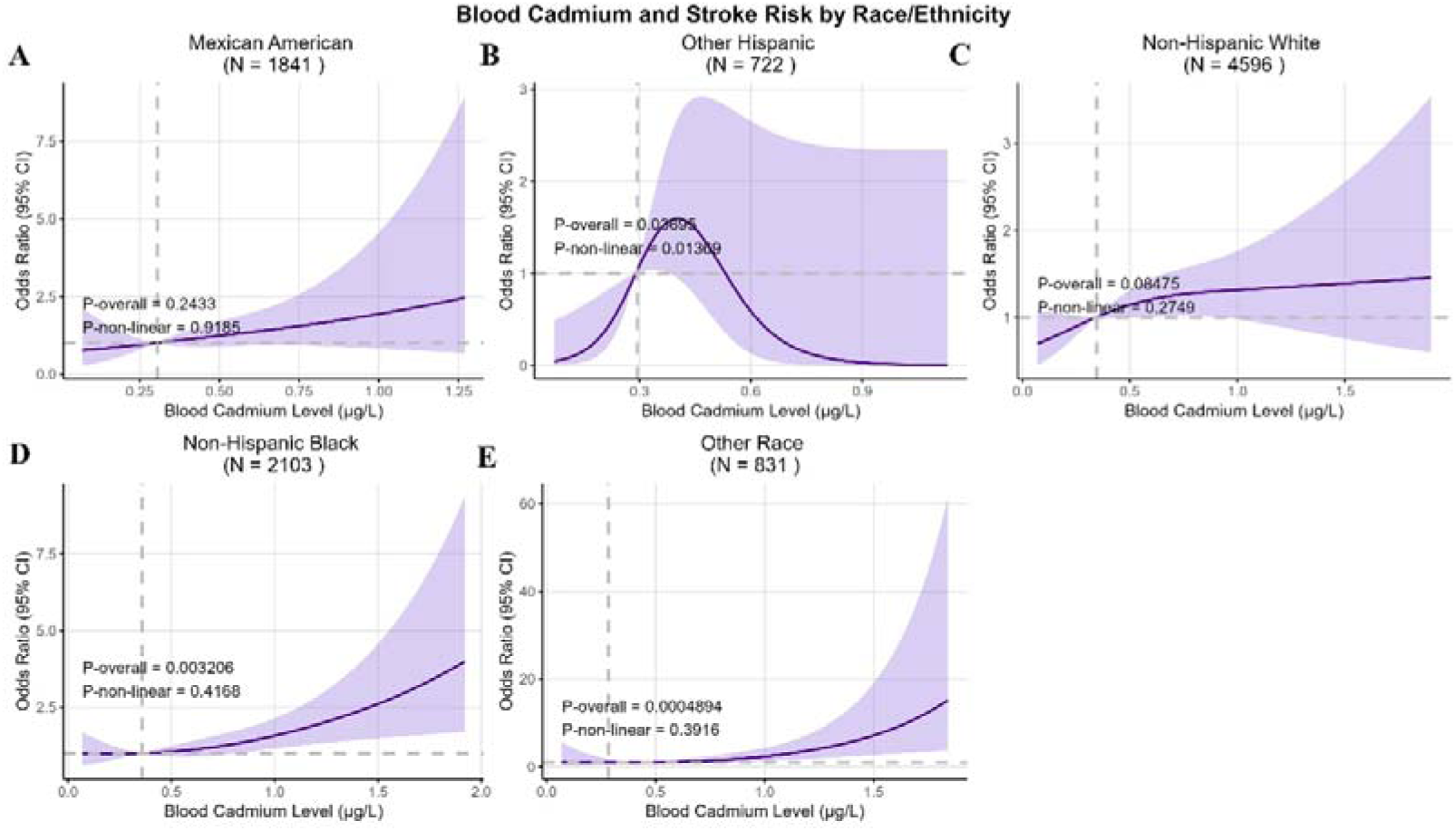
Restricted cubic splines showing dose-response relationships between blood cadmium levels and stroke risk by race/ethnicity. (A) Mexican American (N = 1841), (B) Other Hispanic (N = 722), (C) Non-Hispanic White (N = 4596), (D) Non-Hispanic Black (N = 2103), (E) Other Race (N = 831). Solid lines indicate odds ratios with 95% confidence intervals (shaded areas).

### Subgroup Analysis

Figure 4 presents subgroup analysis results of associations between cadmium exposure and stroke prevalence. Stratified analyses were conducted by age, gender, race/ethnicity, BMI classification, diabetes status, hypertension status, smoking status, alcohol consumption, and physical activity level. In crude models, multiple subgroups showed positive correlations between cadmium exposure and stroke risk, particularly more pronounced associations in populations aged ≥65 years, obese, with diabetes, with hypertension, and low physical activity levels. In adjusted models, association strengths in most subgroups weakened but overall trends remained consistent. Interaction tests for all subgroups did not reach statistical significance (P for interaction > 0.05). (Fig. 4)

**Fig. 4.**
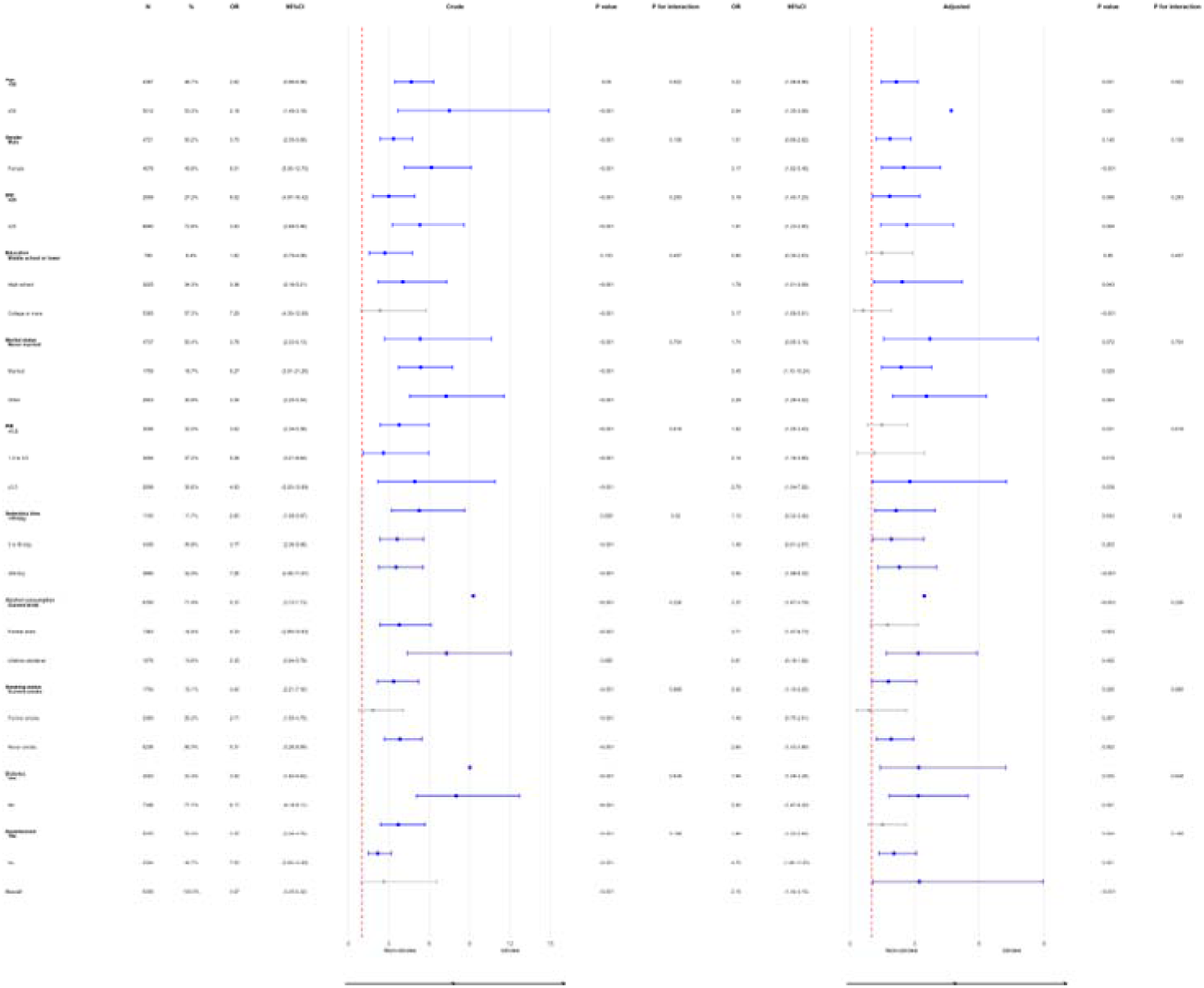
Forest plot of subgroup analyses examining the relationship between blood cadmium exposure and stroke risk stratified by participant characteristics.

### Mediation Effect Analysis

Mediation analysis examined potential roles of lipid and hematological parameters in cadmium exposure and stroke associations. Several biomarkers showed mediation effects. White blood cell count showed the strongest mediation effect, with a mediation proportion of 11.0% (95% CI: 2.1%, 19.9%), reaching statistical significance with a total effect of 0.471 (P < 0.001), indicating that inflammatory responses may be important biological pathways for cadmium exposure affecting stroke risk.

Neutrophil count showed obvious mediation effects, with a mediation proportion of 7.8% (95% CI: 1.4%, 14.2%), reaching statistical significance with a total effect of 0.471 (P < 0.001).

Platelet count mediation proportion was 3.8% (95% CI: 0.6%, 7.0%), reaching statistical significance with a total effect of 0.471 (P < 0.001). HDL cholesterol mediation proportion was 1.5% (95% CI: -0.3%, 3.3%), not reaching statistical significance, with a total effect of 0.471 (P < 0.001).

These results indicate that blood cadmium exposure is associated with stroke risk, with this association being more pronounced in specific population subgroups and primarily operating through biological pathways including inflammation and hematological changes. Identified mediating factors collectively explained approximately 24% of the total effect, with inflammation-related indicators (white blood cell count and neutrophil count) playing major roles. (Fig. 5)

**Fig. 5.**
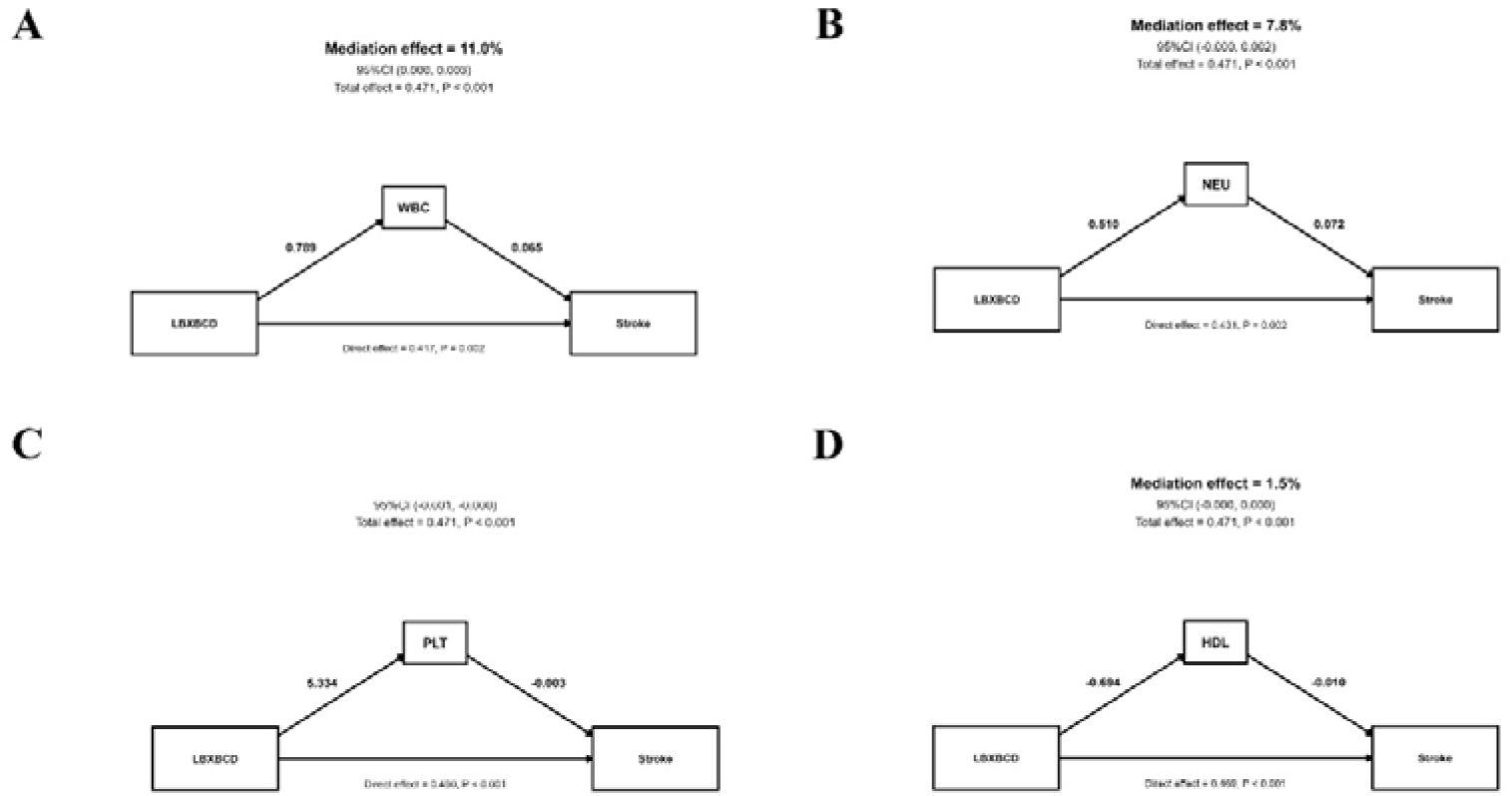
Mediation analysis examining potential biological pathways linking blood cadmium exposure to stroke risk. Path diagrams showing mediation effects of (A) WBC (11.0%), (B) NEU (7.8%), (C) PLT, and (D) HDL (1.5%) in the cadmium-stroke association. WBC, white blood cell count; NEU, neutrophil count; PLT, platelet count; HDL, high-density lipoprotein cholesterol.

## Discussion

This study aimed to explore associations between blood cadmium concentrations and stroke risk using NHANES data. Study results confirmed significant associations between blood cadmium exposure and stroke occurrence, with cadmium as a key environmental toxin driving this association. This study not only confirms existing evidence linking cadmium exposure to adverse cardiovascular outcomes but also provides new insights for understanding cadmium’s independent role in stroke risk. Specifically, our analysis identified cadmium as an important predictor of stroke risk, with each 1 μg/L increase in blood cadmium concentration increasing stroke risk by 48.8%. Importantly, inflammatory responses likely mediate this relationship, as demonstrated by positive correlations between white blood cell counts and cadmium levels, where white blood cell count is an inflammatory marker.

White blood cell count was found to mediate approximately 11.0% of the cadmium-stroke risk association, suggesting that cadmium exposure may increase stroke susceptibility through exacerbating inflammatory pathways.

Contemporary research emphasizes potential health risks from elevated cadmium exposure, positioning such exposure as a contributing factor to adverse cardiovascular outcomes[25]. The role of cadmium exposure as an independent predictor in multifactorial stroke etiology remains a subject of ongoing investigation[26]. Understanding connections between environmental cadmium pollution and stroke is crucial for revealing stroke pathogenesis and guiding public health measures aimed at reducing cadmium exposure, particularly considering cadmium’s widespread distribution as an environmental pollutant and long-term bioaccumulation characteristics[27, 28]. Our study used a large-scale cohort of 10,434 participants to specifically explore associations between blood cadmium concentrations and stroke risk. This study provides new epidemiological evidence for understanding relationships between cadmium, an important environmental toxin, and stroke risk, expanding our understanding of environmental factors’ roles in stroke etiology.

Cadmium, typically inhaled through cigarette smoke, often accumulates in multiple body organs including kidneys, liver, and vascular tissues, with biological half-lives of 10-30 years, potentially leading to a range of cardiovascular health problems including stroke[29]. Oxidative stress and inflammation mechanisms are related to cadmium-induced tissue damage processes, can activate inflammatory signaling pathways, upregulate pro-inflammatory cytokine expression, disrupt vascular endothelial barrier function, and ultimately promote atherosclerosis development[30, 31]. In our analysis, multivariable logistic regression showed highly significant statistical associations between cadmium levels and stroke occurrence.Descriptive analysis showed significant dose-gradient relationships in blood cadmium concentrations among different smoking status populations: daily smokers’ blood cadmium concentrations (1.309±0.987 μg/L) were nearly 4 times higher than never smokers (0.330±0.242 μg/L). More importantly, even in long-term quitters, blood cadmium concentrations (0.419±0.298 μg/L) remained significantly higher than never smokers, reflecting cadmium’s long-term bioaccumulation characteristics in the human body. Based on this finding, we adopted an innovative analysis strategy treating smoking as a mediating variable rather than a traditional confounding factor to more accurately assess cadmium’s independent health effects.

Inflammation and oxidative stress are important components of stroke development mechanisms[32], with research showing they play key roles in stroke pathophysiology. Previous studies have confirmed that chronic inflammatory states promote atherosclerosis development, increase thrombosis risk, and may increase stroke risk by affecting cerebrovascular autoregulation functions[33, 34]. Cadmium exposure can activate inflammatory responses through multiple pathways, including directly activating macrophages and neutrophils, inducing oxidative stress to promote reactive oxygen species production, and disrupting vascular endothelial cell function to increase vascular permeability[35]. Given these shared pathogenic pathways, it is reasonable to consider that cadmium exposure can increase stroke risk through mechanisms involving inflammation and oxidative stress[36].

In our investigation, mediation analysis identified significant positive correlations between inflammatory markers and blood cadmium concentrations. White blood cell count showed the strongest mediation effect with a mediation proportion of 11.0% (95% CI: 2.1%, 19.9%), neutrophil count mediation proportion of 7.8% (95% CI: 1.4%, 14.2%), and platelet count mediation proportion of 3.8% (95% CI: 0.6%, 7.0%), all reaching statistical significance. This finding suggests that higher cadmium exposure may enhance stroke susceptibility by promoting systemic inflammatory states. Identified mediating factors collectively explained approximately 24% of the total effect, with inflammation-related indicators playing major roles.

This study found important effect heterogeneity across different population subgroups, with findings having important clinical and public health significance. Restricted cubic spline analysis showed that in normal triglyceride populations (<150 mg/dL), cadmium exposure showed linear positive correlations with stroke risk (P = 0.008), while in high triglyceride groups (≥200 mg/dL), significant non-linear relationships were observed (P = 0.002, P-non-linear = 0.039). Racial stratification analysis revealed significant population differences, with Non-Hispanic Black populations showing the strongest associations (P = 0.003), possibly reflecting comprehensive influences of genetic background differences, environmental exposure patterns, and socioeconomic factors. These findings provide important evidence for individualized risk assessment and precision prevention strategy development.

This study aimed to explore associations between blood cadmium levels and stroke occurrence within the NHANES framework. Our study results point to significant associations between blood cadmium concentrations and increased stroke incidence, presenting clear dose-response relationships. However, the cross-sectional nature of our study design requires cautious interpretation, as it cannot exclude possibilities of reverse causality. Although cadmium’s long-term bioaccumulation characteristics somewhat mitigate this issue, prospective cohort studies remain the gold standard for establishing causal relationships.

Another limitation is that stroke diagnosis was based on self-reports, which may involve classification bias and cannot distinguish between ischemic and hemorrhagic stroke subtypes. This distinction is crucial because underlying mechanisms, risk factors, and outcomes may differ between the two types. Future research should address these gaps by incorporating stroke subtype data and using longitudinal designs to establish clearer causal pathways.

This study is supported by several important strengths, particularly utilizing innovative statistical methodologies to parse associations between cadmium exposure and stroke risk in large populations. Large sample size (10,434 participants) and nationally representative NHANES data ensure statistical power and generalizability of results. The analysis strategy treating smoking as a mediating variable rather than a confounding factor more accurately assessed cadmium’s independent health effects. Systematic mediation analysis revealed potential biological mechanisms, and detailed stratified analysis identified effect modifiers. This approach not only enhanced the depth of our analysis but also amplified its relevance to clinical practice and public health strategies aimed at mitigating cadmium exposure-related stroke risks.

However, this study also has limitations. Single blood cadmium measurements may not fully reflect long-term exposure status, and possibilities of residual confounding factors or unconsidered variables such as genetic susceptibility remain. While our study identified significant associations between cadmium concentrations and stroke risk, a valuable area for future research would be exploring specific impacts of cadmium exposure on small vessel disease compared to other ischemic stroke subtypes. Future studies should focus on examining these differences to better understand how cadmium exposure affects different stroke mechanisms.

## Conclusions

Cadmium, an important environmental pollutant, has been confirmed to produce harmful effects on the cardiovascular system. The main objective of this study was to examine associations between blood cadmium levels and stroke prevalence using NHANES (1999-2018) data. Our study results suggest that cadmium exposure may lead to increased stroke risk through inflammatory mechanisms, providing new insights for stroke prevention strategies. Specifically, each 1 μg/L increase in blood cadmium concentration increased stroke risk by 48.8%, with the highest exposure quartile showing 118.7% increased stroke risk compared to the lowest quartile.

However, this study’s cross-sectional design limits the ability to establish causal relationships between cadmium exposure and stroke. Future research should focus on longitudinal studies to better understand causal connections between cadmium exposure and stroke development. Additionally, exploring differences in cadmium concentration roles between small vessel disease and other ischemic stroke subtypes, as well as gene-environment interactions that may influence cadmium toxicity susceptibility, are promising directions for future research.

Furthermore, examining interventions and public health policies aimed at reducing cadmium exposure may play key roles in reducing stroke risk and improving overall public health.

## Data Availability

All data produced in the present work are contained in the manuscript.

## Notes

### Competing Interest Statement

The authors have declared no competing interest.

### Funding Statement

This study was supported by the Joint Fund Project of Fujian Provincial Natural Science Foundation in 2022 (No. 2022J011019), the 2022 Fujian Provincial Health and Health Youth Backbone Training Project (No. 2022GA009), the 2024 Fujian Provincial Science and Technology Innovation Joint Fund Project (No. 2024Y907). The funders had no role in the study design, data collection and analysis, decision to publish, or manuscript preparation.

### Author Declarations

The study used (or will use) ONLY openly available human data that were originally located at: https://wwwn.cdc.gov/nchs/nhanes/Default.aspx

